# Mental health of healthcare workers during early phase of COVID19: Variable performance on different factors of stress

**DOI:** 10.1101/2020.09.22.20199323

**Authors:** Seshadri Sekhar Chatterjee, Madhushree Chakrabarty, Shiv Sekhar Chatterjee, Utpal Dan

## Abstract

**Background:** Risks to healthcare workers (HCWs) escalate during pandemics and they are likely to experience a greater level of stress. This cross-sectional study investigated mental distress among HCWs during the early phase of the coronavirus disease-2019 (COVID-19).

**Method:** 140 HCWs of a tertiary care hospital in India were assessed for perceived stress and insomnia. A factor analysis with principal component method reduced these questions to four components which were categorized as insomnia, Stress-related Anxiety, Stress-related Irritability, and Stress-related Hopelessness. Further statistical analyses were done on these factor scores to identify the predictors and investigate the differences between the different categories of HCWs.

**Result:** Doctors were the most anxious among the HCWs. Both doctors and nurses perceived a greater level of irritability than the other HCWs. Compared to the doctors and nurses, other HCWs were more likely to experience insomnia. Lower age, higher education, female gender, and urban habitat were associated with the perception of anxiety. Older age, quarantine, single marital-status predicted irritability. Female gender, single marital-status, and greater ailments contributed to perceived hopelessness. Quarantine significantly predicted insomnia.

**Conclusion:** Different categories of HCWs might experience disparate mental health problems owing to their heterogeneous socio-demographic backgrounds. Customized and personalized care might prove to be helpful in alleviating their problems.

**Highlights:**

1. The doctors and nurses have greater psychological distress than other health-care workers (HCWs).
2. Female HCWs are more likely to have stress related anxiety and hopelessness
3. Insomnia, stress-related anxiety, stress-related irritability, and stress-related hopelessness have different predictive factors.
4. Different categories of HCWs might be affected differently

## 1. Introduction

Coronavirus disease-2019 (COVID-19) has created an unprecedented situation worldwide and has set forth an array of challenges before us — medical, ethical, and organizational ^1^ Health care workers (HCWs) are bound by ethics to provide support to patients ^2^. HCWs across the world are putting their fullest effort to cope with the pandemic and save lives. However, they are not immune to infection. Consequently, HCWs are equally vulnerable to infection as the rest of the population. In fact, the frontline workers are at a greater risk than the general population. Previous statistics clearly indicate that HCWs make a significant portion of the infected cases ^3^.

Owing to increased risk of infection, duty towards patients might tussle with self-preservation and protection of loved ones thereby increasing stress and anxiety of HCWs ^4,5^. Increased duty hours and disrupted biological rhythm during quarantine might lead to insomnia ^6^. Inadequate supply of personal protective equipment, problematic media coverage, and stigma might exacerbate stress ^7–9^. In a recent review of six studies, Spoorthy ^10^ reported that “HCW are encountering a considerable degree of stress, anxiety, depression, insomnia due to the COVID-19 pandemic”. Stress and insomnia are not unitary constructs ^11,12^ rather these two aspects of mental health are intricately intertwined. Research reports that stress and insomnia exacerbate each other and create a vicious cycle impacting long term mental health ^13^. Nonetheless, in certain cases, insomnia might be caused by other factors not related to stress. A data-driven factor analysis can help verify if items of stress and insomnia overlap in this particular sample.

India with its several densely populated states, shortage of medical professionals, inadequate equipment, scarcity of health centers, the paucity of testing facilities, sparse surveillance, and poor awareness among masses, failed to contain the disease^14^. Consequently, the pressure on the health system mounted. The Government of India ordered a nationwide lockdown for 21 days On March 24, 2020. The lockdown was further extended to 31^st^ May with conditional relaxations. The pandemic coupled with lockdown made a deep impact on the socio-economic fabric as well as the mental health conditions of the people. Apprehensions and anguish transformed into fear and stigma towards COVID-19 patients as well as fighters^15^.

Inadequate number of public health care centers along with the escalating COVID-19 treatment expenses in the private health care centers worsened the situation^16^. The already dwindling patient-doctor relationship^17^ reached a worrying level of distrust. Health care workers in general and public health care workers, in particular, suffered acute helplessness. Stigma, work overload, shortage of equipment, dying patients, distrust, concern for personal safety and safety of the family members pushed them into a mental turmoil.

Recent studies on Indian doctors reported significant mental health problems due to COVID-19 ^18–20^. Most of these studies have acquired data through online surveys that have inherent limitations. Lack of focused groups and selection bias threatens the validity of such studies^21^. Here we conducted a pen and paper survey to avoid these limitations^22^.

However, apart from doctors, people working in healthcare facilities such as nurses, ward staff, cleaning staff, porters, and administrative staff are also variably vulnerable 23 and might face mental health problems. People working in certain specialties such as a respiratory ward, infectious diseases ward or critical care ward are subject to greater risk and might be under greater stress.

However, stress is not a unitary construct. It is multifaceted and complex. A global stress score may fail to capture the subtle factors contributing to the elevated stress level. In this study, we conducted a factor analysis on the measures obtained from the perceived stress and the insomnia questionnaires to extract different factors of these two mental health parameters. Subsequently, we investigated if different categories of HCWs have differences in these four different factors of mental health. We also explored if the socio-demographic and the clinical-professional parameters could predict these four factors.

## 2. Material and methods

### 2.1. Ethics

The study was approved by the institutional ethics committee (XXXXX/2020/349/10). All participants signed an informed consent form approved by the above committee.

### 2.2. Settings

The study was conducted from 20th April to 20th May at Diamond Harbour Government Medical College & Hospital (DHGMC), West Bengal, India. During this time COVID-19 was gradually spreading across India thereby mounting pressure on the health care system. DHGMC was converted into a COVID-19 treatment center, well equipped with an isolation ward, quarantine center, fever clinic, and COVID-19 testing facility.

### 2.3. Participants

Of the 550 HCWs employed at DHGMC, 250 randomly selected HCWs, who were on duty during the COVID-19 outbreak, were invited to take part in the study. Participants having any history of neurological or psychiatric illness were excluded from the study. After eliminating participants not meeting the inclusion criteria (n=44), non-responders (n=52), and incomplete data (n=14), finally 140 participants were selected for the study.

After factor analysis, factor scores were scanned for outliers. Three outliers were eliminated from further analyses. Tests for statistical significance were done on 137 participants. These 137 participants were divided into four groups based on their profession. There were 55 doctors (Age: M = 39.22±9.3, 33 Male and 22 Female), 45 nurses (Age: M = 39.60 ±11.6; 0 Male and 45 Female), 20 ward staff (Age: M = 31.45 ±4.8; 20 Male and 0 Female), and 17 non-clinical staff (Age: M = 34.06 ±7.2; 6 Male and 11 Female).

### 2.4. Measures

Demographic information was obtained using a customized demographic data sheet. A questionnaire was designed to assess the participant’s level of exposure to COVID patients. Based on the information they were categorized into four groups — severe risk (specimen collection unit, and isolation ward), high risk (chest/medicine outdoor, fever clinic, and emergency), moderate risk (specialist outpatient and inpatient department), and low risk (administrative work). The **Perceived Stress Scale (**PSS – 10) ^24^ has 10 questions/statements and the respondents indicate their levels of agreement (0 = Never; 1 = Almost; 2 = Sometimes; 3 = Fairly Often 4 = Very Often). It includes items measuring reactions to stressful situations as well as measures of stress. The PSS-10 scale has acceptable reliability measures for Indian population (internal consistency-Cronbach’s α = 0.731; Spearman-Brown split-half reliability coefficient = 0.71)^25^. **Insomnia severity index** (ISI-7) ^26^ contains seven items that assess the severity of both nighttime and daytime components of insomnia. The first three items assess trouble in initiating, maintaining sleep, and early morning awakening. Other items address dissatisfaction with sleep, daytime functions, recognition of insomnia by others, and finally, distress caused by insomnia. These are scored on a five-point scale ranging from 0 = no problem to 4 = very severe problem. The score of 0–7 depicts the absence of insomnia, 8–14 indicates subthreshold insomnia, 15–21 represents moderate, and 22–28 suggests severe insomnia. ISI has high internal consistency (Cronbach’s α = 0.84) test-retest reliability (ICC (2, 1) = 0.84) and validity (correlation with Pittsburgh Sleep Quality Index — r = 0.45) for Indian population^27^. We have used the original English versions of the above tests as all participants in this study had at least 12 years of formal education.

### 2.6. Procedure

Data were collected through a set of self-administered questionnaires, consisting of the above tests. The data were coded and preprocessed in Microsoft Excel (Microsoft Corporation, Washington, USA, 2016). Statistical analyses were performed using Statistical Package for Social Sciences (SPSS) Statistics for Windows, Version 20.0 (IBM Corp., USA, 2011).

### 2.7. Statistical Analyses

A Factor Analysis (FA) using the principal component method with a varimax rotation was conducted on data obtained from 140 patients to reduce the number of variables. We obtained 17 measures per patient: Insomnia (7 questions) and Perceived Stress (10 questions). The KMO value indicated that the sample was factorable (KMO=.768). Homogeneity of variance was confirmed by Bartlett’s test (x^2^ (136) = 926.7, p<. 001). The diagonals of the anti-image correlation matrix were over. 5 for all items except the PES (Q4). This item was dropped from the final analysis. The final factor analysis yielded four factors.

It may be noted here that the factor structure of a particular tool may vary due to sampling differences ^28^. Existing factor analysis data on PSS-10 are based on samples from different cultures and were collected under different socio-economic and health conditions. So, instead of confirmatory factor analysis based on previous studies, a data-driven approach was taken. ‘Eigenvalues greater than one’ was considered as factor extraction criteria since this is considered to be a reliable technique for factor extraction in an exploratory factor analysis ^29^.

Shapiro-Wilk test for normality was done on the total factor scores and it revealed that the data are not normally distributed. After excluding the three outliers (6, 64, and 125) the data conformed to the normality criteria. Hence rest of the analyses were done on these 137 participants.

A mixed-design ANOVA with groups of HCWs (Doctor, N=55; Nurse, N=45; Ward staff, N=20; and Non-Clinical staff, N=17) as a between-subject variable and the four mental health components (Insomnia, Anxiety, Irritability, and Hopelessness) as within-subject variable was conducted to test for an interaction between the groups and the mental health components. This analysis was followed by independent sample t-tests to determine how the groups differed across the four mental health components.

Stepwise regression was conducted to test if socio-demographic (age, gender, habitat, marital-status, education, family, diseases, and media exposure) and clinical-professional variables (duration of service, quarantine, level of risk, contact with confirmed COVID cases, prophylaxis, and use of mask) could predict the mental health components.

## 3. Result

### 3.1. Factor analysis

The final factor analysis was done on 16 items. KMO of the final model was 0.786 and Bartlett’s test was significant ((x^2^ (120) = 877.4, p<0.001) confirming that the data were factorable.^30^ The diagonals of the anti-image correlation matrix were above 0.5 for all items. Communalities were above 0.5 for all items in the final analysis except P-6 (0.42). We extracted four factors with eigenvalues above 1. The four components explained 29.6%, 16.0%, 10.0%, and 6.6% of the variance, respectively. The cumulative percentage of variance explained by the five components was 62.2%. The rotated component matrix with the communalities of the items is given in Table 1.

**Table 1.** Rotated Component Matrix

| Items | Sleeplessness | Anxiety | Irritability | Hopelessness | Communalities |
| --- | --- | --- | --- | --- | --- |
| Insomnia_6 | 0.872 |  |  |  | 0.778 |
| Insomnia_7 | 0.85 |  |  |  | 0.735 |
| Insomnia_5 | 0.792 |  |  |  | 0.697 |
| Insomnia_2 | 0.768 |  |  |  | 0.651 |
| Insomnia_4 | 0.651 |  |  |  | 0.563 |
| Insomnia__1 | 0.618 |  |  |  | 0.522 |
| Insomnia_3 | 0.602 |  |  |  | 0.502 |
| PS_1 |  | 0.835 |  |  | 0.711 |
| PS_3 |  | 0.792 |  |  | 0.654 |
| PS_2 |  | 0.737 |  |  | 0.589 |
| PS_9 |  | 0.721 |  |  | 0.655 |
| PS_5 |  |  | 0.84 |  | 0.697 |
| PS_7 |  |  | 0.74 |  | 0.735 |
| PS_8 |  |  | 0.705 |  | 0.544 |
| PS_10 |  |  |  | 0.725 | 0.59 |
| PS_6 |  |  |  | 0.517 | 0.423 |

Key: PS Perceived stress; INS- Insomnia; SUP-Stress-due-to-unpredictability; SOL-Stress-due-to-overload;; SUC-Stress-due-to-uncontrollability

After scrutinizing the individual items of these four factors, we named them: 1) Insomnia 2) Stress-related Anxiety 3) Stress-related Irritability, and 4) Stress-related Hopelessness. Hereafter these four factors will be referred to as Insomnia, Anxiety, Irritability, and Hopelessness, respectively. Factor *hopelessness* had less than three-item loadings but we retained it as a separate factor because irritability and hopelessness are different aspects of stress. Further analyses were done on these four factor-scores.

### 3.2. Hypothesis testing

The mixed design ANOVA did not yield a significant main effect of mental health factors (F(3, 399) = 0.84, p=.47, observed power =. 24). However, there was a significant main effect of group (F(3, 133) =9.7, p <.001; observed power=.99) and significant Factor scores x Group interaction (F(9, 399) =3.63, p<. 001; observed power =.99). Thus, people from different occupations responded differently to the different mental health factors (Figure1).

**Figure 1.**
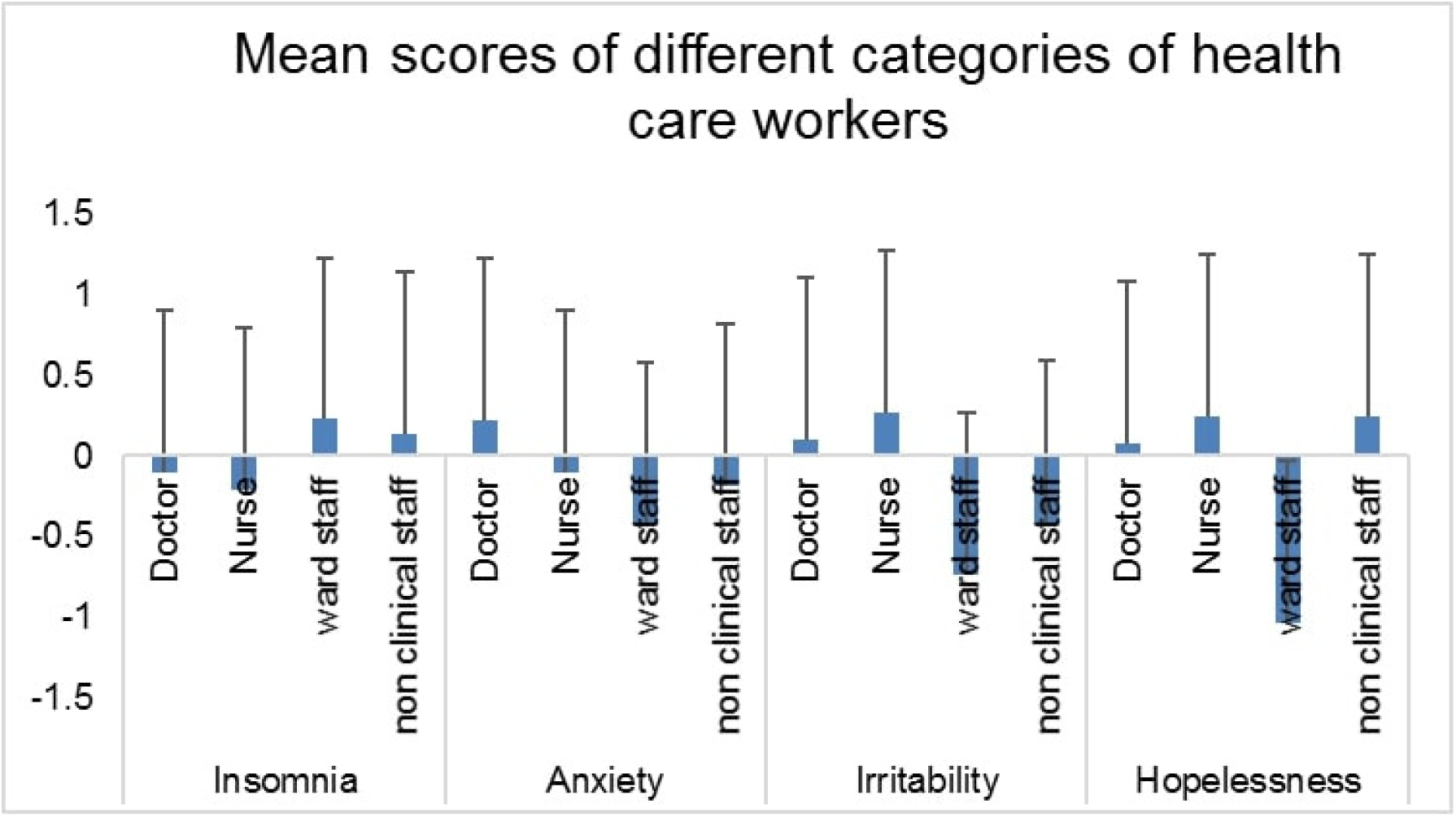
Mean scores of healthcare workers in the four different components of the mental health

Independent sample t-tests revealed that compared to the ward staff, doctors were significantly more anxious, (p =. 005) irritable (p <.001) and hopeless (p =. 001). Nurses were more irritable, (p <.001) and hopeless (p <.001) than the ward staff. Doctors were more irritable than the non-clinical staff (p =.027). Nurses were also more irritable than the non-clinical staff (p =.010).

Nonclinical staff members were more hopeless than the ward staff (p =. 008). Ward staff members experienced more insomnia than the nurses (p =. 01). There were no significant differences between the doctors and the nurses. See Table 2 for details.

**Table 2.** Result of independent sample t-tests

| Factor | Group | Mean | (SD) | Pairs compared | T-value | df | P-value | Cohen's<br>*d* |
| --- | --- | --- | --- | --- | --- | --- | --- | --- |
| Insomnia | Doctor | -0.10 | (1.10) | Doctor - Nurse | 0.528 | 98 | 0.599 | 0.107 |
|  | Nurse | -0.21 | (0.94) | Doctor - WS | -1.923 | 72.989 | 0.058 | 0.399 |
|  | WS | 0.23 | (0.39) | Doctor - NCS | -1.179 | 53.143 | 0.244 | 0.273 |
|  | NCS | 0.14 | (0.57) | Nurse - WS | -2.66 | 62.785 | *0.01 | 0.611 |
|  |  |  |  | Nurse - NCS | -1.422 | 60 | 0.16 | 0.450 |
|  |  |  |  | WS - NCS | 0.578 | 35 | 0.567 | 0.184 |
| Stress-related anxiety | Doctor | 0.22 | (0.83) | Doctor - Nurse | 1.715 | 85.199 | 0.09 | 0.346 |
|  | Nurse | -0.10 | (1.01) | Doctor - WS | 2.917 | 73 | *0.005 | 0.752 |
|  | WS | -0.42 | (0.87) | Doctor - NCS | 1.184 | 19.992 | 0.25 | 0.358 |
|  | NCS | -0.18 | (1.34) | Nurse - WS | 1.23 | 63 | 0.223 | 0.339 |
|  |  |  |  | Nurse - NCS | 0.267 | 60 | 0.79 | 0.067 |
|  |  |  |  | WS - NCS | -0.625 | 26.696 | 0.537 | 0.531 |
| Stress-related irritability | Doctor | 0.10 | (1.05) | Doctor - Nurse | -0.833 | 98 | 0.407 | 0.168 |
|  | Nurse | 0.27 | (0.96) | Doctor - WS | 4.131 | 55.736 | *0.000 | 0.724 |
|  | WS | -0.73 | (0.64) | Doctor - NCS | 2.299 | 39.219 | *<sup>1</sup>0.027 | 0.566 |
|  | NCS | -0.41 | (0.72) | Nurse - WS | 4.939 | 53.303 | *0.000 | 1.22 |
|  |  |  |  | Nurse - NCS | 2.66 | 60 | *0.01 | 0.801 |
|  |  |  |  | WS - NCS | -1.418 | 35 | 0.165 | 0.469 |
| Stress-related hopelessness | Doctor | 0.08 | (0.79) | Doctor - Nurse | -1.224 | 96.425 | 0.224 | 0.232 |
|  | Nurse | 0.24 | (0.57) | Doctor - WS | 3.914 | 25.556 | *0.001 | 1.111 |
|  | WS | -1.03 | (1.17) | Doctor - NCS | -0.418 | 18.623 | 0.681 | 0.129 |
|  | NCS | 0.24 | (1.56) | Nurse - WS | 4.626 | 23.037 | *0.000 | 1.380 |
|  |  |  |  | Nurse - NCS | 0.007 | 17.627 | 0.994 | 0.000 |
|  |  |  |  | WS - NCS | -2.832 | 35 | *0.008 | 0.921 |

Key: WS -Ward staff; NCS – Non-clinical staff; SD - Standard deviation; df - Degree of freedom; \* Statistically significant after FDR correction; \*<sup>1</sup>Statistically not significant after FDR correction

### 3.3. Exploratory analyses

Stepwise linear regression with the socio-demographic variables (age, gender, habitat, marital status, education, family, diseases, and media exposure) as predictors were conducted for all the four factors (Insomnia, anxiety, irritability, and hopelessness). Age (β=, −.431, t = −6.1, p <.001), education (β=. 358, t =4.4, p <.001), gender (β =. 202, t = 2.7,p =. 008), and habitat (β = −.201, t= −2.6,p =. 011) predicted anxiety (F(4,132) =18.27, p <. 001, R^2^ =.356, Cohen’s *f* ^*2*^ =. 552) indicating lower age, higher education, female gender and urban habitat were associated with higher anxiety. Age (β=.480, t = 6.3, p <.001) and marital status (β=.247, t = 3.2, p =. 002) predicted irritability (F (2,134) =22.3, p <. 001, R^2^ =.249, Cohen’s *f*^*2*^ =. 331). Older age and single marital status predicted irritability. Gender (β=. 412, t = 5.2, p <.001), marital status (β= −.203, t = −2.5, p =. 012) and disease (β=. 175, t = 2.3, p =. 025) predicted hopelessness (F(3,133) = 11.4, p <. 001, R^2^ =.205, Cohen’s *f*^*2*^ =. 257). Female gender, single marital status, and greater ailments contributed to perceived hopelessness. None of these variables predicted insomnia.

Stepwise linear regression with the clinical-professional variables (duration of service, quarantine, level of risk, contact with confirmed COVID cases, prophylaxis, and use of mask) as predictors were conducted for all the four factors (insomnia, anxiety, irritability, and hopelessness). Quarantine (β= −.206, t = −2.4, p =. 016) significantly predicted insomnia (F(1,135) = 5.95, p =.016, R^2^ =.042, Cohen’s *f*^*2*^ =. 043). People who were quarantined were more prone to suffer from insomnia. Duration of service (β= −. 467, t = − 5.88, p <. 001) and use of prophylaxis (β= −. 197, t = − 2.5, p =. 015) predicted anxiety (F (2,134) = 17.78, p <.001, R^2^ =.210 Cohen’s *f*^*2*^ =. 265). Fewer years in service and use of prophylaxis was associated with anxiety. Duration of service (β=. 462, t = 6.45, p <.001), quarantine (β= −217, t = − 2.98, p =.003) and level of risk (β= −. 165, t = − 2.3, p =. 024) predicted irritability (F (3,133) = 21.58, p <.001, R^2^ =.327, Cohen’s *f*^*2*^ =. 485). Greater duration of service, quarantine, and a greater level of risk contributed to irritability. None of these variables predicted hopelessness.

## 4. Discussion

Our study aimed to investigate the different components of perceived stress and insomnia experienced by the HCWs and how different socio-demographic and clinical-professional factors influence these components. The factor analysis of insomnia and stress scales yielded four factors which were identified as - 1) Insomnia, 2) Stress-related Anxiety, 3) Stress-related Irritability and 4) Stress-related Hopelessness. The four factors explained 62.2 % of the variance. In this section, we will discuss each of these four factors. Perceived stress yielded three factors and this is consistent with Pangtey et al. ^25^ who validated the Hindi version of PSS-10 in the adult urban population of Delhi.

All the 7 questions of the insomnia scale loaded on the first factor. Insomnia was found to be the most important factor and it explained 29.6% of the variance. There was no significant correlation between the factor insomnia and the three factors of perceived stress. This is consistent with Gupta et al. ^31^ who found no significant difference in perceived stress among three different groups with varying levels of nighttime sleep duration after lockdown due to COVID19. It may be noted that insomnia can be caused by several other factors apart from stress. In this study, quarantine significantly predicted insomnia. More screen time, reduced physical activity, and staying away from home in a quarantine center could contribute to insomnia. Ward staff members were most likely to experience insomnia. Compared to doctors and nurses, other HCWs were more prone to suffer from insomnia.

Stress due to unpredictability has been referred to as *anxiety* in this study. HCWs with lower age, higher education, female gender, and urban habitat experienced higher levels of anxiety. In fact, doctors who formed the most educated group among the HCWs were the most anxious of all. As we have seen in several patients, better knowledge and understanding of the disease can engender stress and anxiety ^32,33^. Doctors are perhaps not an exception to this rule. Female HCWs and HCWs with lower age experienced greater anxiety. This is in line with Matud ^34^ who reported significantly more stress in women even after adjusting for sociodemographic variables. Our result is also in tune with the American Psychological Association’s report of 2019 ^35^ which states that younger adults and women are the more stressed out. This is also partly consistent with Remes et al ^36^ who stated that the prevalence of anxiety disorder is higher in women and young adults. However, it may be noted that anxiety referred here is an aspect of stress and we have not used any tool to measure anxiety per se. Nonetheless, these two psychobiological states are reported to have neural as well as behavioral overlaps ^37^. Our result is consistent with several other studies that report higher levels of stress in people living in cities compared to rural areas ^38,39^. Fewer years in service and use of prophylaxis was associated with anxiety. HCWs with junior titles were probably less adapted to handle such crises and consequently had higher levels of stress. Higher stress levels could result from the use of prophylaxis ^40^. Additionally, people who are more stressed could be more inclined to use prophylaxis.

Stress due to overload has been referred to as *irritability*. Doctors and nurses scored high on this factor compared to other HCWs. This is consistent with recent studies examining the mental health status of HCWs during COVID-19 ^7^. Older and single HCWs were more irritable. This result is quite intuitive. Older people are more likely to succumb to tiredness due to overwork and single HCWs were probably more stressed because they were handling their emotional and physical burden single-handedly. Greater duration of service, quarantine, and a greater level of risk contributed to irritability. This result again is quite expected. Greater duration of service indicates higher age and as already explained older people might capitulate to fatigue and exhaustion more easily than younger people. Moreover, apart from emotional turmoil, quarantine might impose a physical burden as well. Middle-class salaried Indians usually have the privilege of domestic help to take care of household chores. Quarantine could inadvertently repeal this privilege thereby escalating unwonted physical burden and hence stress. This is partly consistent with a study in the general population ^41^ that was covered by the *New Indian Express*. HCWs posted in specialties such as a respiratory ward, infectious- diseases ward, or critical-care ward, where there is a high risk, are plausibly sharing the greatest workload during this pandemic. Consequently, they are probably under greater stress than other HCWs. Wearing the heavy PPE in this hot and humid climate might add to their distress.

Stress due to uncontrollability has been denoted as *hopelessness* in this study. Female gender, single marital-status, and greater ailments contributed to perceived hopelessness. Ward staff members were found to be the most hopeful among the HCWs. Incidentally, all the ward staff members were males. This is in line with the linear regression result that indicated gender as the most important predictor of perceived hopelessness. Female HCWs were more likely to be perturbed with the feeling of hopelessness. Our result is consistent with studies that report a feeling of powerlessness among HCWs. Females, being more empathetic, are perhaps more likely to feel hopeless when they witness people suffering and dying. As in the case of irritability, unmarried HCWs were found to be more hopeless. This is consistent with a recent study that found lower levels of stress hormones in healthy married adults ^42^. Our findings are also in line with Podder et al. ^19^ who reported higher levels of perceived stress in female and unmarried physicians. HCWs with a greater number of ailments had greater perceived hopelessness. Numerous scientific journals and social media platforms are continuously reporting that patients with lung diseases, diabetes, and heart diseases are at increased risk for severe complications from COVID-19. This awareness and a focus on the uncontrollable could worsen the feeling of hopelessness in HCWs with these ailments.^7^.

In sum, this study revealed the fact that HCWs caring for patients during the pandemic are exhibiting significant mental health symptoms. Several factors are contributing to their psychological distress. Perceived Stress is not a unitary construct. It has three different components and these components have different predictive factors. Female HCWs are more likely to have stress-related anxiety and hopelessness. Doctors and nurses have higher levels of stress-related anxiety and irritability. Efforts must be made to provide the much needed mental health care to the HCWs working during the pandemic.

## 5. Conclusion

This data-driven four-factor solution seems to tap insomnia and the three aspects of stress that Cohen (1994) intended to explore —”….how unpredictable, uncontrollable, and overloaded the respondents find their lives”. The study reveals that the HCWs are working with enormous stress and sleep difficulty during the pandemic. Different categories of HCWs are affected differently. Different factors modulate insomnia, stress-related anxiety, stress-related irritability, and stress-related hopelessness.

Small sample size and unequal groups limit the scope of generalizability of the findings of this study. In order to strategically target therapeutic interventions and to establish the possible impact of the pandemic on the mental health of HCWs, confirmation with a larger sample size will be an important next step. The socio-cultural background might influence the intensity and modulate the predictive factors of mental health components. Consequently, while the study from Kashmir ^20^ reports higher levels of stress among male and married HCWs, we found the female and unmarried HCWs more stressed. Future research needs to tease out how social-cultural factors interact with perceived stress. Although this study is preliminary and exploratory in nature, it generates new hypotheses and addresses an important issue that warrants urgent action.

Mental health issues of HCWs need immediate attention. If this goes on unaddressed it might hamper their performance and subsequent health care delivery. This might not only affect their short-term mental wellbeing but could also have a deleterious long term impact ^6,43^. Well-planned pharmacological and non-pharmacological measures must be maneuvered appropriately to grapple the mental health issues of the HCWs in this difficult time. Personalized treatment for different categories of HCWs might help to maximize efficacy.

## Data Availability

Data will be made available on request

## Notes

### Competing Interest Statement

The authors have declared no competing interest.

### Clinical Trial

This is a observational study

### Funding Statement

No funding was received for this study

### Author Declarations

Institutional Ethics Committee, Diamond Harbour Government Medical College and Hospital. Memo No: DHGMC/2020/349/10

